# Antibiotic-resistant bacteria sampled from metropolitan wastewater share genomic similarities with hospital-associated isolates

**DOI:** 10.64898/2025.11.28.25341078

**Authors:** Jakobi T. Deslouches, Nathan J. Raabe, Emma G. Mills, Giuseppe Fleres, Nathan R. Wallace, Mohamed H. Yassin, Daria Van Tyne

## Abstract

Wastewater surveillance is an effective approach for monitoring populations of antibiotic-resistant bacteria and tracking the spread of antimicrobial resistance (AMR) across clinical and environmental settings. In this study, hospital and municipal wastewater were collected monthly for 12 months from hospital and municipal locations in the greater Pittsburgh area to quantify the presence of antibiotic-resistant bacteria and investigate their genomic diversity. After quantitative culturing on six different selective medias, a total of 150 isolates were speciated by 16S rRNA sequencing, which revealed diverse pathogenic and non-pathogenic taxa, including *Klebsiella* spp., *Pseudomonas* spp. and *Aeromonas* spp. A subset of isolates belonging to clinically relevant species (n = 46) underwent whole-genome sequencing, which identified several antibiotic resistance genes of clinical concern, such as *bla*_KPC_, *bla*_NDM_, and *bla*_IMP_, and revealed genetic similarities between wastewater and clinical isolates collected from infected patients at a Pittsburgh-area medical center. In addition, analysis of plasmids carried by wastewater isolates revealed closely related plasmids present in isolates from different species and sampling locations. Overall, these findings demonstrate that both hospital and municipal wastewater act as interconnected reservoirs of antimicrobial resistance. Integrating wastewater surveillance with clinical and genomic data could enable the early detection of emerging resistance threats and support proactive infection-control strategies.

**IMPORTANCE:** Antibiotic-resistant bacteria persist within hospital environments and cause deadly infections. This study examined the prevalence of antibiotic-resistant bacteria in both hospital and municipal wastewater sampled from the Pittsburgh metropolitan area. Over a 12-month surveillance period, we observed a high abundance of antibiotic-resistant organisms, including pathogenic Gram-positive and Gram-negative species encoding diverse antibiotic resistance genes, many of which were plasmid-encoded. We also identified instances of closely related isolates sampled from wastewater and clinical sources, as well as closely related plasmids found in different species and sampled from different wastewater sites. Overall, this study demonstrates that wastewater surveillance is a viable method for non-invasive monitoring of resistant bacteria at a local scale.

## INTRODUCTION

Bacteria are developing resistance to antibiotics at an alarming rate, with many strains exhibiting resistance to multiple classes of antibiotics (1). These resistant organisms pose a burden to healthcare systems by increasing morbidity, mortality, and healthcare costs (1,2). Many antibiotic-resistant infections are caused by a group known as ESKAPE pathogens (*Enterococcus* spp., *Staphylococcus aureus, Klebsiella pneumoniae*, *Acinetobacter baumannii*, *Pseudomonas aeruginosa*, and *Enterobacter* spp.), which includes pathogens encoding multiple strategies for developing resistance and evading therapeutics (3). Most ESKAPE pathogens develop resistance through horizontal gene transfer (HGT), which involves the sharing of mobile genetic elements (MGEs) such as plasmids, transposons, prophages, and insertion sequences (4–6). Multidrug resistant ESKAPE pathogens cause hospital-acquired infections which are often difficult to treat and have high mortality rates (3,7).

Microbial surveillance is the monitoring of microorganisms present in a specific area or region over time. Surveillance of microorganisms can aid in epidemiologic studies, including point-prevalence surveys and outbreak investigations (8). Wastewater surveillance is a form of microbial surveillance that samples water from a sewer system adjacent to a community and is sometimes more feasible than obtaining biospecimens directly from infected patients (9). Wastewater surveillance may also capture a larger population of organisms, as it aggregates sampling across an entire municipality or catchment area. Furthermore, wastewater systems harbor high microbial diversity, providing an environment where HGT frequently occurs among bacteria (10). Wastewater surveillance was used to monitor the prevalence of SARS-CoV-2 during the COVID-19 pandemic, and has been used in the past to monitor the presence and prevalence of antibiotic resistance genes (9,11). The use of whole genome sequencing (WGS) provides additional opportunities for comparative analyses between antibiotic-resistant bacteria sampled from wastewater and those collected from clinical settings (12).

Here we combined wastewater surveillance and WGS to quantify and characterize antibiotic-resistant bacteria sampled from hospital and municipal wastewater sources in the Pittsburgh area over a 12-month period. We performed quantitative plating, species typing, WGS, and comparative genomics analysis to understand the prevalence and diversity of antibiotic-resistant bacteria sampled from both hospital and municipal environments. We also compared the genomes of antibiotic-resistant bacteria isolated from wastewater with those of clinical isolates collected from a local hospital system through the Enhanced Detection System for Healthcare-Associated Transmission (EDS-HAT) project (13,14). Our results reveal a high abundance of antibiotic-resistant bacteria across both hospital and municipal wastewater sites, including isolates encoding antibiotic resistance genes of high concern.

## METHODS

### Wastewater sampling

Hospital and municipal wastewater samples were collected concurrently and approximately monthly over a 12-month period (2023-2024) from four different sampling locations (two hospital and two municipal) in the Pittsburgh area on a concurrent basis. On the day of collection, wastewater samples from each site were serially diluted 10-fold in 1x Phosphate Buffer Saline (PBS) and 10uL of each dilution was plated onto selective media agars. Selective plates used included MacConkey agar (MAC) containing 1 µg/mL ciprofloxacin (CIP) for detection of fluoroquinolone-resistant Enterobacterales, MacConkey agar (MAC) containing 1 µg/mL cefotaxime (CTX) for detection of ESBL-producing Enterobacterales, MacConkey agar (MAC) containing 1 µg/mL meropenem (MEM) for detection of carbapenem-resistant Enterobacterales, Pseudomonas isolation agar (PIA) containing 1 µg/mL meropenem (MEM) for detection of carbapenem-resistant *Pseudomonas spp.*, mannitol salt agar (MSA) containing 4 µg/mL oxacillin (OXA) for detection of methicillin-resistant *Staphylococcus spp*., and bile esculin azide agar (BEA) containing 10 ug/mL vancomycin (VAN) for the detection of vancomycin-resistant *Enterococcus* spp. (VRE). After overnight incubation at 37°C, colony-forming units (CFU) per mL were calculated and recorded for each collection site and timepoint by plate type.

### 16S rRNA sequencing

Seven different sampling time points were selected for isolation of individual colonies from selective media plates. Between 1-3 individual colonies with different morphologies (150 isolates total) were picked from selective plates and passaged on Brain Heart Infusion (BHI) agar with no antibiotic selection to create monocultures. Plates were scraped and isolates were cryopreserved in 1 mL BHI media + 50% glycerol and stored at −80°C. 16S rRNA typing was performed by Azenta Life Sciences (Burlington, MA) using PCR assays to type the V1 to V9 regions of the bacterial 16S rRNA gene.

### Whole genome sequencing (WGS) and comparative genomics analysis

A total of 46 antibiotic-resistant wastewater isolates belonging to clinically relevant bacterial species were selected to undergo WGS. DNA from pure overnight cultures grown in tryptic soy broth (TSB) media at 37°C shaking at 170 rpm was extracted using a Qiagen DNeasy Blood and Tissue Kit following the manufacturer’s instructions. After quantification using a Qubit fluorimeter, next-generation sequencing libraries were prepared and sequenced using 2 x 150 paired-end reads on an Illumina NextSeq at SeqCenter (Pittsburgh, PA). Kraken2 v2.1.2 was used for initial taxonomic classification of all WGS samples (15). Reads were assembled into contigs (≥500 bp) using SPAdes v3.15.5 (16), and QUAST v5.2.0 (17) was used to assess the quality of each assembled genome. To verify taxonomic classifications, Kleborate v2.4.1 (18) was used for *Klebsiella* spp. and GTDB-Tk v2.4.1 (19) was used for other genera. Prokka v1.14.5 (20) was used to annotate all genomes, while GTDB-Tk v2.4.1 was used to generate a core genome alignment and RAxML v8.2.12 (21) was run using the GTRCAT algorithm with 100 bootstraps to generate a phylogenetic tree for subsequent visualization in iTOL v7.2 (22). AMRFinderPlus v4.0.22 (23) was used to screen for the presence of antibiotic resistance genes, using the – organism flag for the most closely related species in the database and an identity threshold of ≥ 90%. The genetic relatedness of wastewater isolate genomes was determined using whole genome single nucleotide polymorphisms (SNPs) identified via Split Kmer Analysis (SKA) (24). Multilocus sequence types (STs) were identified using the PubMLST database with mlst v2.11 (25,26). Genomes were also compared to previously collected hospital isolates deposited in NCBI BioProject PRJNA475751 using SKA and a 20 SNP cut-off for clustering of genetically similar isolates.

### Long read sequencing and hybrid assembly

A total of 19 isolates were selected for long-read sequencing on the Oxford Nanopore Technology MinION platform. DNA from pure overnight cultures grown in TSB at 37°C shaking at 170 rpm was extracted using a Qiagen DNeasy Blood and Tissue Kit following the manufacturer’s instructions. Sequencing libraries were prepared using the SQK-RBK114.24 rapid gDNA Barcoding Kit and were sequenced for 72 hours using an Oxford Nanopore Technologies (ONT) MinION Mk1C on R10.4.1 flow cells. Basecalling and demultiplexing were performed using Dorado v0.9.6. Hybrid assembly was performed using both short-read (Illumina) and long-read (ONT) data for each isolate with Unicycler v0.5.1 (27), and *de novo* assemblies were visualized using Bandage v0_8_1 (28). QUAST was used to screen for contamination, verify genome sizes, and perform other quality control checks on each hybrid assembly (17).

### Plasmid analysis

Plasmids which met the following inclusion criteria were identified and extracted from hybrid assemblies: circular sequence, ≥10kb in length, and a plasmid incompatibility group and/or replicon identified by mob_typer using default parameters (29). For plasmids where a generic replicon group was assigned (*i.e.,* “rep_cluster”), the corresponding sequence from the mob_typer database was queried using BLASTp against the PlasmidFinder database (30) using >80% similarity score (% coverage x % identity) to identify the closest replicon type (31). Using these criteria, a total of 57 plasmid contigs were extracted from hybrid assemblies across 19 antibiotic-resistant wastewater isolates. ABRicate v1.0.1 was used to identify antibiotic resistance genes within plasmids using the Resfinder database (32) with ≥80% cutoffs for percent identity and coverage. Pling v2.0.0 (33) was used to quantify plasmid similarity and perform clustering of highly related plasmids into communities (containment distance <0.3) and subcommunities (additionally, DCJ-indel distance ≤4). Clusters of similar plasmids were visualized using Gephi v0.10 (34). Clustered plasmids were annotated using Prokka v1.14.5 (20) and aligned with Easyfig v2.2.5 to generate multiple sequence alignments (35).

## RESULTS

### Population dynamics of resistant organisms over time

To assess the abundance of antibiotic-resistant bacteria present in hospital and municipal wastewater, we sampled four different sites (two hospital, two municipal) located in the Pittsburgh, Pennsylvania metropolitan area monthly over a one-year period and used selective media plating to quantify resistant organism burdens across each site and over time. Temporal monitoring showed variable dynamics across each wastewater source (**Figure 1, Table S1**). Overall, plating of wastewater onto MacConkey agar containing ciprofloxacin, cefotaxime, or meropenem, as well as Pseudomonas isolation agar with meropenem, revealed an abundance of growth, with 10^4^-10^5^ colony-forming units (CFU) per mL detected on average year-round. The abundance of growth on these plates from hospital wastewater was more variable month-to-month compared to municipal wastewater, which showed less variability between sampling time points. Plating of wastewater onto mannitol salt agar with oxacillin and bile esculin azide agar with vancomycin yielded far lower CFU/mL burdens compared to the other selective medias, with some hospital sampling timepoints showing no growth above the limit of detection. Plating on Gram-negative–selective medias yielded 2.03 × 10^5^ CFU/mL on average, whereas plating on Gram-positive–selective medias yielded 1.2 × 10^3^ CFU/mL on average, representing a nearly 170-fold difference in abundance (**Table S1**). Additionally, the abundance of organisms from municipal sources that grew on these plates was seasonally variable, with lower abundances apparent in months six through nine. We also compared organism burdens on each selective media between sampling locations, and observed modest but significant differences in growth between the two municipal sites on MacConkey agar with ciprofloxacin and on bile esculin azide agar with vancomycin (**Figure S1**). Overall, these data suggest that antibiotic-resistant organisms were detected at moderate abundance in both hospital and municipal wastewater, and only modest differences in abundance were observed between different sample locations and times.

**Figure 1:**
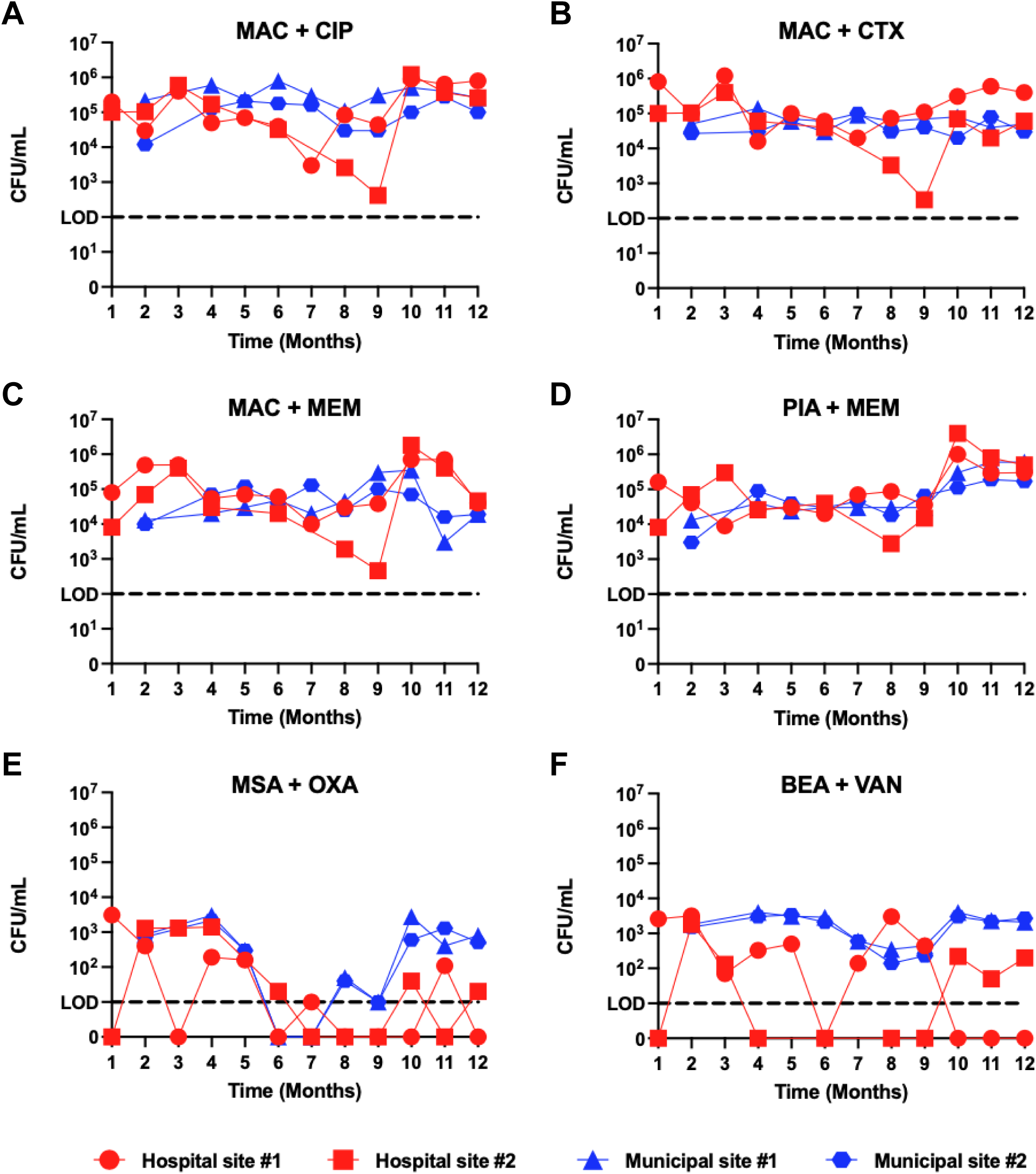
Temporal quantification of resistant organism burdens from hospital and municipal wastewater samples grown on selective medias. Tracking of colony-forming units (CFU) per mL over time for four different wastewater sources sampled over a 12-month period by plating onto six different selective medias, including A) MacConkey agar (MAC) containing 1 µg/mL ciprofloxacin (CIP), B) MAC containing 1 µg/mL cefotaxime (CTX), C) MAC containing 1 µg/mL meropenem (MEM), D) Pseudomonas isolation agar (PIA) containing 1 µg/mL MEM, E) Mannitol salt agar (MSA) containing 4 µg/mL oxacillin (OXA), and F) Bile esculin azide agar (BEA) containing 10ug/mL vancomycin (VAN). Colors and symbol shapes distinguish hospital and municipal sites, and lines connect data points from the same sampling site.

### 16S rRNA typing of wastewater isolates

To assess the taxonomic distribution of organisms that grew on the selective medias we used, we performed 16S rRNA typing on 150 isolates sampled from the four different sites at seven different sampling timepoints. While a wide variety of organisms were identified at each site, the most abundant genera observed across the dataset included *Aeromonas* spp. (n = 37 isolates, 25%), *Klebsiella* spp. (n= 28 isolates, 19%), *Pseudomonas* spp. (n = 20 isolates, 13%), and *Enterococcus* spp. (n = 15 isolates, 10%) (**Figure 2, Table S2**). These genera were identified at all four sites, while *Staphylococcus* spp., *Raoultella* spp., and *Citrobacter* spp. were identified at three of the four sites. There was no apparent enrichment of any genera at any site, suggesting that bacterial communities across the four locations that we sampled were largely comprised of similar taxa.

**Figure 2:**
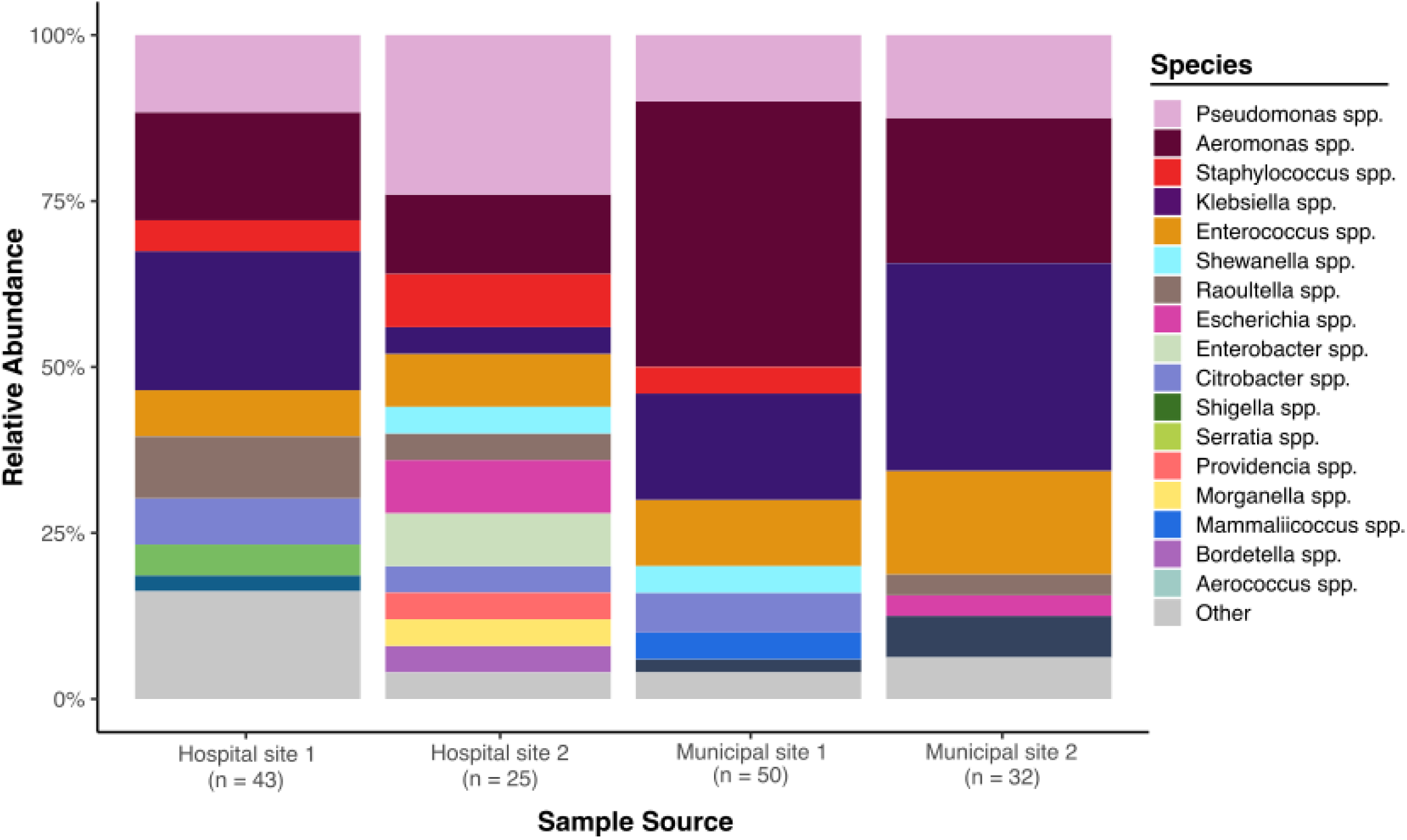
Relative abundance of bacterial genera isolated from hospital and municipal wastewater. Isolated colonies (n = 150 total) collected from wastewater samples plated on selective medias were typed via 16S rRNA sequencing. The relative abundance of isolates belonging to each genus from each sampling location is shown.

### Genomic analysis of wastewater isolates

The 16S rRNA typing analysis we performed identified several species belonging to the ESKAPE group of pathogens as well as other pathogenic Enterobacterales such as *Citrobacter* spp., *Raoultella* spp., and *Serratia* spp. To investigate the genomic diversity of isolates belonging to these pathogen groups, we performed whole genome sequencing of 46 antibiotic-resistant wastewater isolates on the Illumina platform and constructed a maximum likelihood phylogenetic tree based on an alignment of 120 conserved bacterial core genes (**Figure 3**). Isolates were broadly divided into Gram-positive (n = 12) and Gram-negative (n = 34) groups. Species identification revealed a diverse population of organisms, with *Klebsiella* spp. (n = 18, 39%), *Enterococcus* spp. (n = 13, 28%), and *Raoultella* spp. (n = 6, 13%) being the most frequently observed (**Table S3**). Species identification from genomic data largely agreed with 16S typing results, with the exception of isolates within the *Klebsiella pneumoniae* species complex for which WGS showed greater accuracy in species identification.

**Figure 3:**
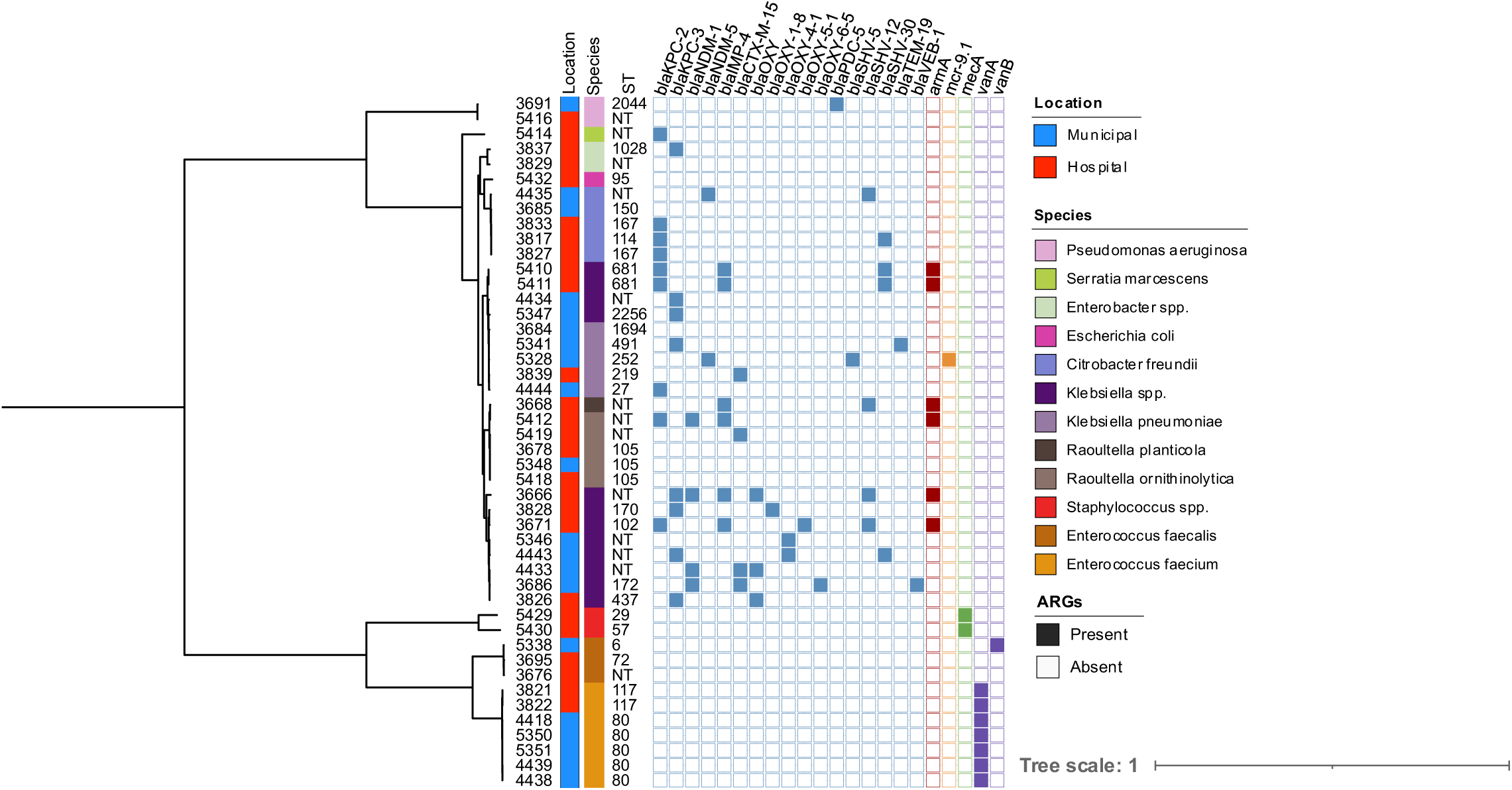
Genetic relatedness and presence of high-risk antimicrobial resistance genes among 46 isolates collected from wastewater sources. The midpoint-rooted phylogenetic tree was constructed using RAxML with 100 bootstraps based on a 120 bacterial marker gene alignment produced by GTDB-Tk. The location where isolate was collected from is shown as municipal (blue) or hospital (red) wastewater sources. Species group are colored as indicated. Multilocus sequence types are noted, those non-typable are indicated as “NT”. Antimicrobial resistance genes were identified using AMRFinderPlus with gene presence noted by a filled square. Genes are color based on expected phenotypic resistance: aminoglycoside (red), β-lactams (light blue), colistin (orange), methicillin, (green), and glycopeptide (purple).

To assess for the presence of antibiotic resistance genes among sequenced isolates, isolate genomes were queried against the AMRFinderPlus database. This analysis revealed that most wastewater isolates possessed genes predicted to confer resistance to multiple antibiotic classes. (**Figure 3, Table S3**). The most frequently observed classes of antibiotic resistance genes included aminoglycosides, β-lactams, and quinolones, with different resistance patterns apparent between the different genera sampled. Of the 34 Gram-negative isolates sequenced, 22 isolates (65%) were carbapenemase-positive, most commonly due to the presence of a *bla*_KPC_ (n = 17, 77%), *bla*_NDM_ (n = 6, 27%), and/or *bla*_IMP-4_ (n = 6, 27%). Seven isolates encoded more than one carbapenemase, with two isolates collected from hospital wastewater carrying three different enzymes. Six isolates from diverse *Klebsiella* (n = 4) and *Raoultella* species (n = 2) encoded the *armA* 16S methyltransferase, which is predicted to confer resistance to all clinically relevant aminoglycosides (36). Concerningly, these isolates were also carbapenemase-positive. Additionally, of all sequenced isolates, 39% (n = 18) encoded extended-spectrum β-lactamases (ESBLs), including a variety of *bla*_CTX,_ *bla*_OXY,_ and *bla*_SHV_ enzymes. A single *Klebsiella pneumoniae* isolate encoded the colistin resistance gene *mcr-9.1* (37) in addition to the *bla*_NDM-5_ carbapenemase and the *bla*_CTX-M-15_ ESBL. We identified two *E. faecalis* isolates that grew on vancomycin-supplemented media despite lacking a detectable van operon, suggesting breakthrough growth or loss of the van operon during laboratory propagation prior to WGS. Nonetheless, our results indicate that high-risk antimicrobial resistance genes are not limited to healthcare settings, as they were detected in both municipal and hospital wastewater samples.

### Genetic relatedness of wastewater and clinical isolates

To assess the genetic relatedness of antibiotic-resistant wastewater isolates between each other and on a larger epidemiological scale, we quantified the number of single nucleotide polymorphisms (SNPs) between sequenced isolates sampled from wastewater as well as between wastewater isolates and clinical isolates collected from infected patients at the University of Pittsburgh Medical Center (UPMC) (**Figure 4**). Three pairs of *E. faecium* isolates sampled from wastewater were found to be closely related to one another (0-15 SNPs apart). These isolates were collected from the same sampling location and same time point, suggesting that they represent strains that were abundant in the water that was sampled. We also identified three clinical isolates (2 *E. faecium* and 1 *K. pneumoniae*) that were closely related to wastewater isolates (7-15 SNPs apart). In two instances, the wastewater isolates were collected from hospital wastewater, although the hospitals where the wastewater was collected differed from those where the patients were sampled. The three *E. faecium* clusters contained isolates belonging to pandemic lineages ST80 and ST117, which are increasing in prevalence both locally and globally (38,39) (**Table S3**). Taken together, these data suggest that most wastewater isolates in our study were unrelated to one another. However, the instances of genetic relatedness between wastewater and clinical samples demonstrate the potential for spread of resistant organisms between clinical and environmental settings.

**Figure 4:**
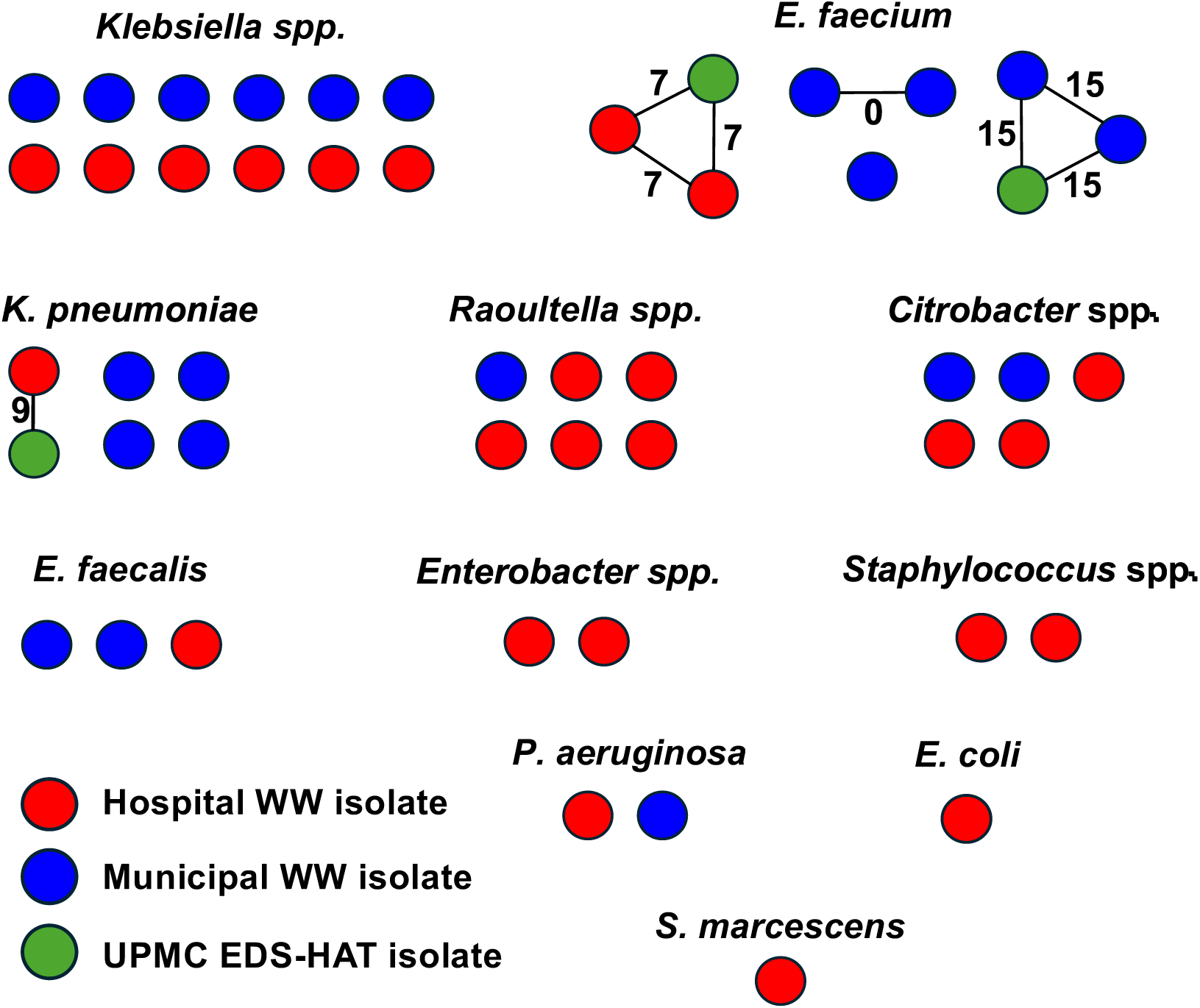
Cluster network plot of genetic relatedness between isolates sampled from wastewater and clinical isolates. Red circles represent hospital wastewater isolates, blue circles represent municipal wastewater isolates, and green circles represent clinical isolates collected from the Enhanced Detection System for Healthcare-Associated Transmission (EDS-HAT) program. Isolates are grouped by species. Sold lines connect isolates that are closely related (i.e. ≤15 single nucleotide polymorphisms, SNPs, apart). SNP distances between clustered isolate pairs are listed next to each line.

### Genetically similar plasmids identified in hospital and municipal wastewater isolates

Of the 46 sequenced wastewater isolates, we selected 19 isolates for MinION sequencing, performed hybrid genome assembly, and extracted a total of 57 circular plasmids greater than 10kb in length with an identifiable plasmid incompatibility group and/or replicon type from the resulting genomes (**Table S4**). Among these 57 plasmids we identified 16 distinct incompatibility groups and/or replicon types, most commonly IncFII (n = 27, 47%), IncFIB (n = 13, 23%), IncM1 (n = 4, 7%), and IncU (n = 4, 7%). Many of these plasmids encoded antibiotic resistance genes, some of which confer resistance to multiple classes of antibiotics, namely β-lactams, aminoglycosides, and macrolides. Clustering of plasmid sequences using Pling (33) identified four clusters of genetically similar plasmids with distinct incompatibility/replicon types containing two to four plasmids each: IncFII (n=4), IncM1 (n=4), IncU/repFIB/repHI5B (n=2), and IncX3 (n=2) (**Figure 5A, Table S4**). The two largest clusters (IncFII and IncM1) contained plasmids from different species and were recovered from different sampling locations, including both hospital and municipal wastewater sources. Within a cluster, plasmids demonstrated reasonably high homology between shared sequences, but the presence of antibiotic resistance genes varied between plasmids (**Figure 5B**). Recovery of highly similar plasmids from isolates originating from both hospital and municipal wastewater sources suggests that some plasmids may circulate between these wastewater systems and could transfer antibiotic resistance and other genes between bacteria in these settings.

**Figure 5:**
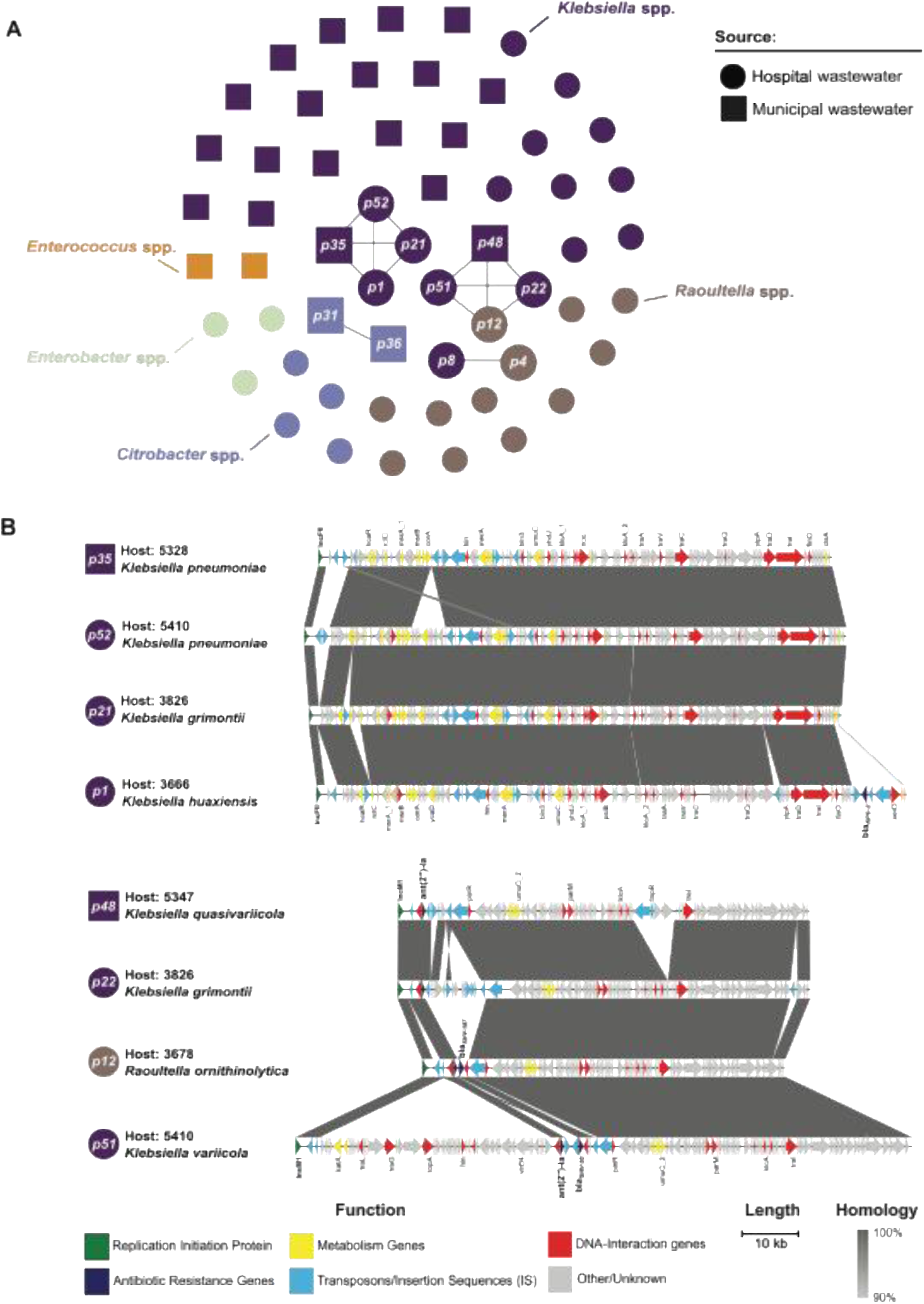
Genetic relatedness of plasmids encoded by antibiotic-resistant wastewater isolates. A) Cluster plot of 57 extracted plasmids from wastewater isolates subjected to MinION sequencing. Plasmids are grouped by species group and genetically similar plasmids (double-cut-and-join value ≤4, containment <0.3 identified by Pling) are connected with lines. B) Multiple sequence alignments of two clusters of genetically similar plasmids. Grey shading indicates genetically similar sequences (≥90% identity, ≥500 bp length). Open reading frames are colored based on gene annotation, and plasmids are labeled as in (A).

## DISCUSSION

This study monitored the occurrence of antibiotic-resistant bacteria sampled from hospital and municipal wastewater across the greater Pittsburgh area, and characterized the genomic variability and antibiotic resistance gene content of nearly 50 individual isolates sampled from wastewater. By combining longitudinal sampling, selective culturing, whole-genome sequencing, and comparative analysis of bacterial and plasmid sequences, we captured both the abundance and diversity of antibiotic-resistant taxa, and resolved the genetic contexts of their antimicrobial-resistance determinants. Comparisons with contemporaneous clinical isolates from a hospital in our region revealed closely related isolates that were shared between wastewater and patient populations, suggesting possible overlap between environmental and clinical reservoirs.

Similar to prior studies, we identified a diverse array of antibiotic-resistant bacterial species, most commonly *Aeromonas* spp., *Klebsiella* spp., *Pseudomonas* spp., and *Enterococcus* spp. within both hospital and municipal wastewater settings (40–47). *Klebsiella* spp., *Pseudomonas* spp., and *Enterococcus* spp. are opportunistic pathogens with antibiotic resistance primarily driven by gene acquisition or chromosomal alterations (48–50). *Aeromonas* spp. are considered environmental and nonpathogenic to humans; however, they can cause opportunistic infections and are intrinsically resistant to carbapenems due to a chromosomally encoded metallo-β-lactamase genes (51). Previous wastewater surveillance studies using PCR-based methods have reported carbapenemase profiles similar to those observed here, including *bla*_KPC_, *bla*_NDM_, and *bla*_IMP_ variants (42,52,53). However, by using whole-genome sequencing, here we identified both the presence of these resistance determinants and their distribution across bacterial genera in both hospital and community wastewater. This comparative genomic approach provides greater insight into the mechanisms driving antimicrobial resistance dissemination within interconnected environmental and healthcare reservoirs.

To investigate potential connections between environmental and clinical reservoirs of antimicrobial resistance, we compared the genomes of wastewater isolates with each other and with patient isolates collected through a local healthcare-associated transmission surveillance program (13). This analysis revealed three putative clusters consisting of both wastewater and clinical isolate genomes, suggesting possible overlap between clinical and environmental settings. Notably, the clustered *E. faecium* isolates belonged to globally disseminated lineages ST117 and ST80, both of which are associated with hospital-adapted clades responsible for outbreaks and persistent colonization in healthcare systems (38,53,54). The detection of these epidemic lineages in wastewater aligns with prior studies showing wastewater as a reservoir for healthcare-associated pathogens (55,56). Furthermore, *E. faecium* and *K. pneumoniae* are the most frequent causes of hospital outbreaks at our healthcare center (14,57,58), which aligns with our observation of closely related isolates of these species also detected in wastewater. While most isolates in our collection were genetically distinct from one another, the small number of highly related isolates we identified shows the importance of genomic surveillance to monitor the spread of antibiotic-resistant pathogens in environmental and community settings.

Through comparing the sequences of plasmids encoded by the isolates we sampled, we identified four clusters of closely related plasmids present in diverse isolates sampled from municipal and hospital wastewater sources. These clusters all contained conjugative plasmids, which can readily transfer between bacterial hosts and facilitate the spread of antimicrobial resistance elements across both hospital and municipal wastewater environments (59–61). Consistent with previous reports, our findings show plasmids as a likely driver of antimicrobial resistance propagation within diverse bacterial populations and environmental reservoirs (59,62). Notably, we identified two isolates carrying an IncX3 plasmid encoding *bla*_NDM-5_ which were both collected from municipal wastewater. IncX3 plasmids harboring *bla*_NDM-5_ have been widely reported across multiple countries and isolated from various sources including hospitalized patients, agriculture settings, and wastewater (63–66). Our findings, in conjunction with other reports (59–61,63), highlight wastewater as a potential reservoir for plasmid-mediated dissemination of high-risk resistance genes, showing the need for continued environmental surveillance.

While our approach uncovered key genomic similarities between environmental and clinical antibiotic-resistant bacteria, some limitations should be acknowledged. First, our analysis prioritized sampling depth over breadth, focusing on four specific sewer lines within the greater Pittsburgh area and therefore limiting broader geographic inferences. Second, only a subset of clinically relevant genera underwent whole-genome sequencing, leaving non-clinical genera such as *Aeromonas* and *Shewanella* excluded from downstream analyses despite their recognized roles in antimicrobial resistance dissemination (67–69). In addition, the use of culture-based methods inherently biased our detection toward organisms capable of growth under the conditions we employed, which overlooked viable but non-culturable, slow-growing, anaerobic, and low-abundance taxa. Metagenomic sequencing could provide a more comprehensive view of the bacterial community and resistome composition in these samples, and this will be a focus of our future work. Finally, monthly sampling intervals may have missed short-term fluctuations in bacterial abundance or horizontal gene transfer events.

In summary, this study demonstrates that hospital and municipal wastewater function as interconnected reservoirs for antibiotic-resistant bacteria and plasmids of high clinical relevance. By integrating a culture-based approach with whole-genome sequencing and comparative genomics analysis, we found that environmental and healthcare niches within a single metropolitan area shared overlapping pools of strains, plasmids, and antimicrobial resistance genes. These findings underscore the importance of wastewater genomic surveillance as a complementary and non-invasive tool for tracking high-risk resistance determinants and guiding infection-prevention strategies. Integrating environmental and clinical genomic data in real time will be essential for the early detection of emerging resistance threats and for developing targeted interventions to curb their dissemination. Ultimately, linking environmental and clinical surveillance data could strengthen antimicrobial stewardship efforts, aid responses to outbreaks of antibiotic-resistant bacteria, and inform public health policies aimed at containing the spread of antibiotic resistance.

## SUPPLEMENTAL MATERIAL

**TableS1.** Bacterial counts (CFU/mL) in wastewater across selective media and timepoints.

**TableS2.** Species identification by 16S rRNA sequencing.

**TableS3.** Antimicrobial resistance genes detected in sequenced wastewater isolates.

**TableS4.** Circular plasmids identified from hybrid genome assemblies of wastewater isolates.

**Figure S1. Quantification of resistant organism burdens from hospital and municipal wastewater.** Plots display colony-forming units (CFU) per mL of wastewater sourced from four different locations. Selective medias included: A) MacConkey agar (MAC) containing 1 µg/mL ciprofloxacin (CIP), B) MAC containing 1 µg/mL cefotaxime (CTX), C) MAC containing 1 µg/mL meropenem (MEM), D) Pseudomonas isolation agar (PIA) containing 1 µg/mL MEM, E) Mannitol salt agar (MSA) containing 4 µg/mL oxacillin (OXA), and F) Bile esculin azide agar (BEA) containing 10ug/mL vancomycin (VAN). Pairwise comparisons between CFU/mL at each location were performed via Mann-Whitney test adjusted for multiple comparisons (L < 0.0083); p < 0.0001: ****, p = 0.0005: ***, p = 0.0036: **.

## DATA AVAILABILITY

Raw Illumina sequencing reads and plasmid sequences are uploaded to NCBI Sequence Read Archive (SRA) and GenBank under BioProject PRJNA1311597. All other data is provided in the supplemental material accompanying the manuscript.

## ACKNOWLEDGEMENTS

We gratefully acknowledge all members of the Van Tyne lab for their helpful input throughout the course of this study. The study was supported by the Department of Medicine at the University of Pittsburgh School of Medicine. The funders had no role in study design, data collection and analysis, decision to publish, or preparation of the manuscript.

## REFERENCES

1. Centers for Disease Control and Prevention (U.S.). Antibiotic resistance threats in the United States, 2019 [Internet]. Centers for Disease Control and Prevention (U.S.); 2019 Nov [cited 2025 June 27]. Available from: https://stacks.cdc.gov/view/cdc/82532

2. Poudel AN, Zhu S, Cooper N, Little P, Tarrant C, Hickman M, et al. The economic burden of antibiotic resistance: A systematic review and meta-analysis. Karunasagar I, editor. PLoS ONE. 2023 May 8;18(5):e0285170.

3. Santajit S, Indrawattana N. Mechanisms of Antimicrobial Resistance in ESKAPE Pathogens. BioMed Research International. 2016;2016:1–8.

4. De Oliveira DMP, Forde BM, Kidd TJ, Harris PNA, Schembri MA, Beatson SA, et al. Antimicrobial Resistance in ESKAPE Pathogens. Clin Microbiol Rev. 2020 June 17;33(3):e00181–19.

5. Maguire M, Serna C, Montero Serra N, Kovarova A, O’Connor L, Cahill N, et al. Spatiotemporal and genomic analysis of carbapenem resistance elements in Enterobacterales from hospital inpatients and natural water ecosystems of an Irish city. Van Tyne D, editor. Microbiol Spectr [Internet]. 2025 Jan 7 [cited 2025 July 11];13(1). Available from: https://journals.asm.org/doi/10.1128/spectrum.00904-24

6. Conte D, Mesa D, Jové T, Zamparette CP, Sincero TCM, Palmeiro JK, et al. Novel Insights into *bla*_GES_ Mobilome Reveal Extensive Genetic Variation in Hospital Effluents. Tyne DV, editor. Microbiol Spectr [Internet]. 2022 Aug 31 [cited 2025 July 11];10(4). Available from: https://journals.asm.org/doi/10.1128/spectrum.02469-21

7. Davidova-Gerzova L, Lausova J, Sukkar I, Nesporova K, Nechutna L, Vlkova K, et al. Hospital and community wastewater as a source of multidrug-resistant ESBL-producing Escherichia coli. Front Cell Infect Microbiol. 2023;13:1184081.

8. Galvin S, Dolan A, Cahill O, Daniels S, Humphreys H. Microbial monitoring of the hospital environment: why and how? Journal of Hospital Infection. 2012 Nov;82(3):143–51.

9. Augusto MR, Claro ICM, Siqueira AK, Sousa GS, Caldereiro CR, Duran AFA, et al. Sampling strategies for wastewater surveillance: Evaluating the variability of SARS-COV-2 RNA concentration in composite and grab samples. J Environ Chem Eng. 2022 June;10(3):107478.

10. Tiwari A, Kurittu P, Al-Mustapha AI, Heljanko V, Johansson V, Thakali O, et al. Wastewater surveillance of antibiotic-resistant bacterial pathogens: A systematic review. Front Microbiol. 2022 Dec 15;13:977106.

11. Johar AA, Salih MA, Abdelrahman HA, Al Mana H, Hadi HA, Eltai NO. Wastewater-based epidemiology for tracking bacterial diversity and antibiotic resistance in COVID-19 isolation hospitals in Qatar. Journal of Hospital Infection. 2023 Nov;141:209–20.

12. Mehra R, Meda M, Pichon B, Gentry V, Smith A, Nicholls M, et al. Whole-genome sequencing links cases dispersed in time, place, and person while supporting healthcare worker management in an outbreak of Panton–Valentine leucocidin meticillin-resistant Staphylococcus aureus; and a review of literature. Journal of Hospital Infection. 2023 Nov;141:88–98.

13. Sundermann AJ, Chen J, Kumar P, Ayres AM, Cho ST, Ezeonwuka C, et al. Whole-Genome Sequencing Surveillance and Machine Learning of the Electronic Health Record for Enhanced Healthcare Outbreak Detection. Clin Infect Dis. 2022 Aug 31;75(3):476–82.

14. Sundermann AJ, Kumar P, Griffith MP, Waggle KD, Rangachar Srinivasa V, Raabe N, et al. Real-Time Genomic Surveillance for Enhanced Healthcare Outbreak Detection and Control: Clinical and Economic Impact. Clin Infect Dis. 2025 Apr 30;ciaf216.

15. Wood DE, Lu J, Langmead B. Improved metagenomic analysis with Kraken 2. Genome Biol. 2019 Nov 28;20(1):257.

16. Prjibelski A, Antipov D, Meleshko D, Lapidus A, Korobeynikov A. Using SPAdes De Novo Assembler. CP in Bioinformatics. 2020 June;70(1):e102.

17. Gurevich A, Saveliev V, Vyahhi N, Tesler G. QUAST: quality assessment tool for genome assemblies. Bioinformatics. 2013 Apr 15;29(8):1072–5.

18. Lam MMC, Wick RR, Watts SC, Cerdeira LT, Wyres KL, Holt KE. A genomic surveillance framework and genotyping tool for Klebsiella pneumoniae and its related species complex. Nat Commun. 2021 July 7;12(1):4188.

19. Chaumeil PA, Mussig AJ, Hugenholtz P, Parks DH. GTDB-Tk: a toolkit to classify genomes with the Genome Taxonomy Database. Hancock J, editor. Bioinformatics. 2020 Mar 1;36(6):1925–7.

20. Seemann T. Prokka: rapid prokaryotic genome annotation. Bioinformatics. 2014 July 15;30(14):2068–9.

21. Stamatakis A. RAxML version 8: a tool for phylogenetic analysis and post-analysis of large phylogenies. Bioinformatics. 2014 May 1;30(9):1312–3.

22. Letunic I, Bork P. Interactive Tree Of Life (iTOL) v5: an online tool for phylogenetic tree display and annotation. Nucleic Acids Research. 2021 July 2;49(W1):W293–6.

23. Feldgarden M, Brover V, Gonzalez-Escalona N, Frye JG, Haendiges J, Haft DH, et al. AMRFinderPlus and the Reference Gene Catalog facilitate examination of the genomic links among antimicrobial resistance, stress response, and virulence. Sci Rep. 2021 June 16;11(1):12728.

24. Harris SR. SKA: Split Kmer Analysis Toolkit for Bacterial Genomic Epidemiology [Internet]. Genomics; 2018 [cited 2025 June 30]. Available from: http://biorxiv.org/lookup/doi/10.1101/453142

25. Seemann T. mlst [Internet]. Github; Available from: https://github.com/tseemann/mlst

26. Jolley KA, Maiden MC. BIGSdb: Scalable analysis of bacterial genome variation at the population level. BMC Bioinformatics. 2010 Dec;11(1):595.

27. Wick RR, Judd LM, Gorrie CL, Holt KE. Unicycler: Resolving bacterial genome assemblies from short and long sequencing reads. Phillippy AM, editor. PLoS Comput Biol. 2017 June 8;13(6):e1005595.

28. Wick RR, Schultz MB, Zobel J, Holt KE. Bandage: interactive visualization of *de novo* genome assemblies. Bioinformatics. 2015 Oct 15;31(20):3350–2.

29. Robertson J, Nash JHE. MOB-suite: software tools for clustering, reconstruction and typing of plasmids from draft assemblies. Microb Genom. 2018 Aug;4(8):e000206.

30. Carattoli A, Hasman H. PlasmidFinder and In Silico pMLST: Identification and Typing of Plasmid Replicons in Whole-Genome Sequencing (WGS). Methods Mol Biol. 2020;2075:285–94.

31. Carattoli A, Zankari E, García-Fernández A, Voldby Larsen M, Lund O, Villa L, et al. In silico detection and typing of plasmids using PlasmidFinder and plasmid multilocus sequence typing. Antimicrob Agents Chemother. 2014 July;58(7):3895–903.

32. Florensa AF, Kaas RS, Clausen PTLC, Aytan-Aktug D, Aarestrup FM. ResFinder - an open online resource for identification of antimicrobial resistance genes in next-generation sequencing data and prediction of phenotypes from genotypes. Microb Genom. 2022 Jan;8(1):000748.

33. Frolova D, Lima L, Roberts L, Bohnenkämper L, Wittler R, Stoye J, et al. Applying rearrangement distances to enable plasmid epidemiology with pling [Internet]. Bioinformatics; 2024 [cited 2025 June 27]. Available from: http://biorxiv.org/lookup/doi/10.1101/2024.06.12.598623

34. Bastian M, Heymann S, Jacomy M. Gephi: An Open Source Software for Exploring and Manipulating Networks. ICWSM. 2009 Mar 19;3(1):361–2.

35. Sullivan MJ, Petty NK, Beatson SA. Easyfig: a genome comparison visualizer. Bioinformatics. 2011 Apr 1;27(7):1009–10.

36. Doi Y, Wachino JI, Arakawa Y. Aminoglycoside Resistance: The Emergence of Acquired 16S Ribosomal RNA Methyltransferases. Infect Dis Clin North Am. 2016 June;30(2):523–37.

37. Song K, Jin L, Cai M, Wang Q, Wu X, Wang S, et al. Decoding the origins, spread, and global risks of mcr-9 gene. EBioMedicine. 2024 Oct;108:105326.

38. Mills EG, Hewlett K, Smith AB, Griffith MP, Pless L, Sundermann AJ, et al. Bacteriocin production facilitates nosocomial emergence of vancomycin-resistant Enterococcus faecium. Nat Microbiol. 2025 Apr;10(4):871–81.

39. Egan SA, Kavanagh NL, Shore AC, Mollerup S, Samaniego Castruita JA, O’Connell B, et al. Genomic analysis of 600 vancomycin-resistant Enterococcus faecium reveals a high prevalence of ST80 and spread of similar vanA regions via IS1216E and plasmid transfer in diverse genetic lineages in Ireland. J Antimicrob Chemother. 2022 Feb 1;77(2):320–30.

40. Punch R, Azani R, Ellison C, Majury A, Hynds PD, Payne SJ, et al. The surveillance of antimicrobial resistance in wastewater from a one health perspective: A global scoping and temporal review (2014-2024). One Health. 2025 Dec;21:101139.

41. Tiwari A, Paakkanen J, Österblad M, Kirveskari J, Hendriksen RS, Heikinheimo A. Wastewater Surveillance Detected Carbapenemase Enzymes in Clinically Relevant Gram-Negative Bacteria in Helsinki, Finland; 2011-2012. Front Microbiol. 2022;13:887888.

42. Pereira AL, de Oliveira PM, Faria-Junior C, Alves EG, de Castro E Caldo Lima GR, da Costa Lamounier TA, et al. Environmental spreading of clinically relevant carbapenem-resistant gram-negative bacilli: the occurrence of blaKPC-or-NDM strains relates to local hospital activities. BMC Microbiol. 2022 Jan 4;22(1):6.

43. Blaak H, Kemper MA, de Man H, van Leuken JPG, Schijven JF, van Passel MWJ, et al. Nationwide surveillance reveals frequent detection of carbapenemase-producing *Enterobacterales* in Dutch municipal wastewater. Science of The Total Environment. 2021 July 1;776:145925.

44. Magalhães MJTL, Pontes G, Serra PT, Balieiro A, Castro D, Pieri FA, et al. Multidrug resistant Pseudomonas aeruginosa survey in a stream receiving effluents from ineffective wastewater hospital plants. BMC Microbiology. 2016 Aug 24;16(1):193.

45. Zhang Q, Zhang S, Xu B, Dong L, Zhao Z, Li B. Molecular Epidemiological Characteristics of Carbapenem Resistant Aeromonas from Hospital Wastewater. Infect Drug Resist. 2024;17:2439–48.

46. Urzua-Abad MM, Aquino-Andrade A, Castelan-Vega JA, Merida-Vieyra J, Ribas-Aparicio RM, Belmont-Monroy L, et al. Detection of carbapenemases in Enterobacterales and other Gram-negative bacilli recovered from hospital and municipal wastewater in Mexico City. Sci Rep. 2024 Nov 4;14(1):26576.

47. Radisic V, Grevskott DH, Lunestad BT, Øvreås L, Marathe NP. Sewage-based surveillance shows presence of *Klebsiella pneumoniae* resistant against last resort antibiotics in the population in Bergen, Norway. International Journal of Hygiene and Environmental Health. 2023 Mar 1;248:114075.

48. Miller WR, Munita JM, Arias CA. Mechanisms of antibiotic resistance in enterococci. Expert Rev Anti Infect Ther. 2014 Oct;12(10):1221–36.

49. Zheng S, Li S, Zhang D, Zhang X, Zhou D, Hou Q, et al. The mechanisms of antibiotic resistance and drug resistance transmission of Klebsiella pneumoniae. J Antibiot. 2025 Nov;78(12):704–16.

50. Meletis G, Exindari M, Vavatsi N, Sofianou D, Diza E. Mechanisms responsible for the emergence of carbapenem resistance in Pseudomonas aeruginosa. Hippokratia. 2012 Oct;16(4):303–7.

51. Segatore B, Massidda O, Satta G, Setacci D, Amicosante G. High specificity of cphA-encoded metallo-beta-lactamase from Aeromonas hydrophila AE036 for carbapenems and its contribution to beta-lactam resistance. Antimicrob Agents Chemother. 1993 June;37(6):1324–8.

52. Mollenkopf DF, Lee S, Ballash GA, Sulliván SMP, Lee J, Wittum TE. Carbapenemase-producing Enterobacterales and their carbapenemase genes are stably recovered across the wastewater-watershed ecosystem nexus. Science of The Total Environment. 2025 May 1;975:179241.

53. Teban-Man A, Szekeres E, Fang P, Klümper U, Hegedus A, Baricz A, et al. Municipal Wastewaters Carry Important Carbapenemase Genes Independent of Hospital Input and Can Mirror Clinical Resistance Patterns. Microbiology Spectrum. 2022 Mar 2;10(2):e02711–21.

54. Lebreton F, van Schaik W, McGuire AM, Godfrey P, Griggs A, Mazumdar V, et al. Emergence of epidemic multidrug-resistant Enterococcus faecium from animal and commensal strains. mBio. 2013 Aug 20;4(4):e00534–13.

55. Chaudhury S, Oransathit W, Peerapongpaisarn D, Oransathit W, Thamnurak C, Pradipol C, et al. Identification of clinically relevant multi-drug resistant ESKAPEE isolates from hospital wastewater surveillance in Thailand. Front Microbiol. 2025;16:1657219.

56. Mahnert A, Moissl-Eichinger C, Zojer M, Bogumil D, Mizrahi I, Rattei T, et al. Man-made microbial resistances in built environments. Nat Commun. 2019 Feb 27;10(1):968.

57. Marsh JW, Mustapha MM, Griffith MP, Evans DR, Ezeonwuka C, Pasculle AW, et al. Evolution of Outbreak-Causing Carbapenem-Resistant Klebsiella pneumoniae ST258 at a Tertiary Care Hospital over 8 Years. mBio. 2019 Sept 3;10(5):e01945–19.

58. Sundermann AJ, Rangachar Srinivasa V, Mills EG, Griffith MP, Evans E, Chen J, et al. Genomic sequencing surveillance of patients colonized with vancomycin-resistant enterococci improves detection of healthcare-associated transmission. BMC Glob Public Health. 2025 Aug 13;3(1):69.

59. Smyth C, Leigh RJ, Do TT, Walsh F. Communities of plasmids as strategies for antimicrobial resistance gene survival in wastewater treatment plant effluent. npj Antimicrob Resist. 2025 Sept 9;3(1):78.

60. Dmowski M, Gołębiewski M, Kern-Zdanowicz I. Characteristics of the Conjugative Transfer System of the IncM Plasmid pCTX-M3 and Identification of Its Putative Regulators. Journal of Bacteriology. 2018 Aug 24;200(18):10.1128/jb.00234-18.

61. Lin D, Xie M, Li R, Chen K, Chan EWC, Chen S. IncFII Conjugative Plasmid-Mediated Transmission of blaNDM-1 Elements among Animal-Borne Escherichia coli Strains. Antimicrobial Agents and Chemotherapy. 2016 Dec 27;61(1):10.1128/aac.02285-16.

62. Risely A, Newbury A, Stalder T, Simmons BI, Top EM, Buckling A, et al. Host-plasmid network structure in wastewater is linked to antimicrobial resistance genes. Nat Commun. 2024 Jan 16;15(1):555.

63. Hammer-Dedet F, Aujoulat F, Jumas-Bilak E, Licznar-Fajardo P. Persistence and Dissemination Capacities of a blaNDM-5-Harboring IncX-3 Plasmid in Escherichia coli Isolated from an Urban River in Montpellier, France. Antibiotics. 2022 Feb;11(2):196.

64. Ho PL, Wang Y, Liu MCJ, Lai ELY, Law PYT, Cao H, et al. IncX3 Epidemic Plasmid Carrying blaNDM-5 in Escherichia coli from Swine in Multiple Geographic Areas in China. Antimicrob Agents Chemother. 2018 Feb 23;62(3):e02295–17.

65. Tian D, Wang B, Zhang H, Pan F, Wang C, Shi Y, et al. Dissemination of the blaNDM-5 Gene via IncX3-Type Plasmid among Enterobacteriaceae in Children. mSphere. 2020 Jan 8;5(1):e00699–19.

66. Raabe NJ, Valek AL, Griffith MP, Mills E, Waggle K, Srinivasa VR, et al. Real-time genomic epidemiologic investigation of a multispecies plasmid-associated hospital outbreak of NDM-5-producing Enterobacterales infections. Int J Infect Dis. 2024 May;142:106971.

67. Dong N, Zhang Y, Wu Y, Ju X, Yan Z, Liu C, et al. Genetic insights into Shewanella spp., progenitor of the blaOXA-48-like genes: a large-scale study. Microbial Genomics. 2025;11(6):001417.

68. Lee HJ, Storesund JE, Lunestad BT, Hoel S, Lerfall J, Jakobsen AN. Whole genome sequence analysis of Aeromonas spp. isolated from ready-to-eat seafood: antimicrobial resistance and virulence factors. Front Microbiol [Internet]. 2023 June 30 [cited 2025 Oct 29];14. Available from: https://www.frontiersin.org/journals/microbiology/articles/10.3389/fmicb.2023.1175304/full

69. Dhanapala PM, Kalupahana RS, Kalupahana AW, Wijesekera DPH, Kottawatta SA, Jayasekera NK, et al. Characterization and Antimicrobial Resistance of Environmental and Clinical Aeromonas Species Isolated from Fresh Water Ornamental Fish and Associated Farming Environment in Sri Lanka. Microorganisms. 2021 Oct;9(10):2106.

