## Supplementary material for "Antibiotic-resistant bacteria sampled from metropolitan wastewater share genomic similarities with hospital-associated isolates": Figure S1

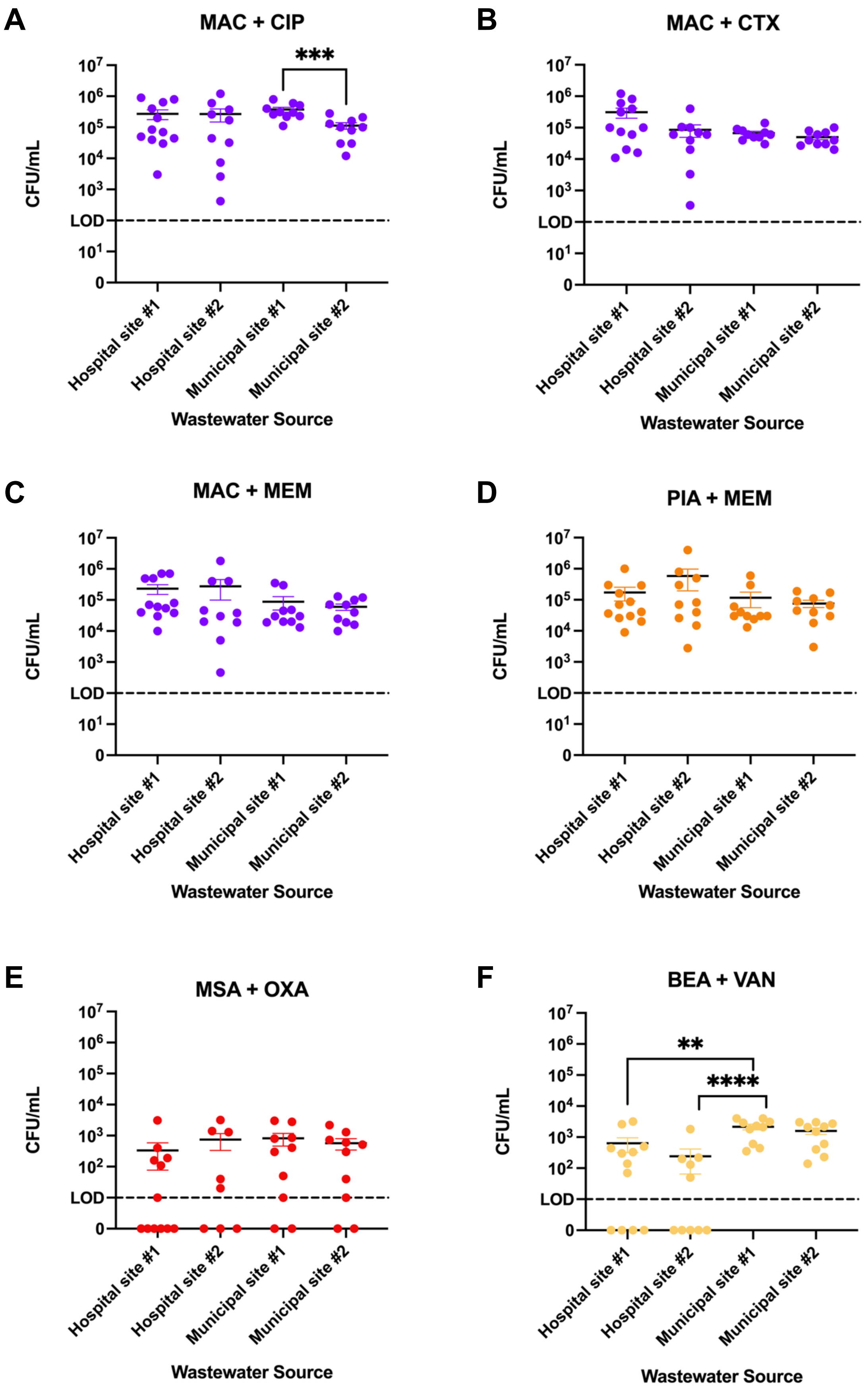


**Figure S1: Quantification of resistant organism burdens from hospital and municipal wastewater.** Plots display colony-forming units (CFU) per mL of wastewater sourced from four different locations. Selective medias included: A) MacConkey agar (MAC) containing 1 µg/mL ciprofloxacin (CIP), B) MAC containing 1 µg/mL cefotaxime (CTX), C) MAC containing 1 µg/mL meropenem (MEM), D) Pseudomonas isolation agar (PIA) containing 1 µg/mL MEM, E) Mannitol salt agar (MSA) containing 4 µg/mL oxacillin (OXA), and F) Bile esculin azide agar (BEA) containing 10ug/mL vancomycin (VAN). Pairwise comparisons between CFU/mL at each location were performed via Mann-Whitney test adjusted for multiple comparisons ($\alpha < 0.0083$); $p < 0.0001$: ****, $p = 0.0005$: ***, $p = 0.0036$: **.
